# Soluble angiotensin-converting enzyme 2 as a prognostic biomarker for disease progression in patients infected with SARS-CoV-2

**DOI:** 10.1101/2021.10.13.21264901

**Authors:** Noelia Diaz Troyano, Pablo Gabriel Medina, Stephen Weber, Martin Klammer, Raquel Barquin-DelPino, Laura Castillo-Ribelles, Angels Esteban, Manuel Hernández-González, Roser Ferrer-Costa, Tomas Pumarola, Francisco Rodriguez Frias

## Abstract

**Background:** There is a need for better prediction of disease severity in patients infected with severe acute respiratory syndrome coronavirus 2 (SARS-CoV-2). Soluble angiotensin-converting enzyme 2 (sACE2) arises from shedding of membrane ACE2 (mACE2) that is known to be a receptor for the spike protein of SARS-CoV-2; however, its value as a biomarker for disease severity is unknown. This study evaluated the predictive value of sACE2 in the context of other known biomarkers of inflammation and tissue damage (C-reactive protein [CRP], growth/differentiation factor-15 [GDF-15], interleukin-6 [IL-6], and soluble fms-like tyrosine kinase-1 [sFlt-1]) in patients with and without SARS-CoV-2 with different clinical outcomes.

**Methods:** For univariate analyses, median differences between biomarker levels were calculated for the following patient groups classified according to clinical outcome: reverse transcription polymerase chain reaction (RT-PCR)-confirmed SARS-CoV-2 positive (Groups 1–4); RT-PCR-confirmed SARS-CoV-2 negative following previous SARS-CoV-2 infection (Groups 5 and 6); and RT-PCR-confirmed SARS-CoV-2 negative controls (Group 7).

**Results:** Median levels of CRP, GDF-15, IL-6, and sFlt-1 were significantly higher in patients with SARS-CoV-2 who were admitted to hospital compared with patients who were discharged (all p<0.001), whereas levels of sACE2 were significantly lower (p<0.001). Receiver operating characteristic curve analysis of sACE2 provided cut-offs for the prediction of hospital admission of ≤0.05 ng/mL (positive predictive value: 89.1%) and ≥0.42 ng/mL (negative predictive value: 84.0%).

**Conclusion:** These findings support further investigation of sACE2, either as a single biomarker or as part of a panel, to predict hospitalisation risk and disease severity in patients infected with SARS-CoV-2.

**HIGHLIGHTS:** Noelia Diaz Troyano: Noy-Lee-ah Dee-az Tro-yah-no

Better prediction of disease severity in patients with severe acute respiratory syndrome coronavirus 2 (SARS-CoV-2) is needed. We measured soluble angiotensin-converting enzyme 2 (soluble ACE2) and other biomarkers of inflammation and tissue damage in patients recruited from Vall d’Hebron University Hospital, with and without SARS-CoV-2 and with different clinical outcomes. Levels of soluble ACE2 were significantly lower in patients with SARS-CoV-2 who had the most severe clinical outcome in all comparisons. These findings support a protective role for soluble ACE2 in SARS-CoV-2 infection and warrant further investigation of soluble ACE2 as a biomarker for disease severity in patients with SARS-CoV-2.

## INTRODUCTION

Severe acute respiratory syndrome coronavirus 2 (SARS-CoV-2) is the causal agent of coronavirus disease 2019 (COVID-19), the global outbreak of which was declared a pandemic on 11 March 2020 by the World Health Organisation (1). To date, over 4 million deaths have been recorded due to COVID-19 (2). The virus predominantly targets the respiratory system, initially causing local virus-mediated tissue damage, followed by a second phase in which infected host cells trigger an immune response. In severe SARS-CoV-2 infection, the activation of the immune system results in a cytokine storm, causing a local and systemic inflammatory response, and leading to dysfunction and damage in other organ systems (3). Any proposed biomarker algorithm for SARS-CoV-2 disease progression should therefore include markers of inflammation and tissue damage.

Membrane-bound angiotensin-converting enzyme 2 (mACE2) serves as a receptor for the spike protein of SARS-CoV-2 (4) and is also a part of the renin angiotensin system (RAS). In cardiovascular disease, the classical arm of the RAS system is commonly dysregulated and exerts the majority of its pathological effects through the peptide hormone angiotensin II. In contrast, ACE2 and its products angiotensin-(1–7) and angiotensin-(1–9) form the counter-regulatory arm of the RAS system, and have protective effects at the pulmonary (anti-fibrotic) and cardiovascular (vasodilator) levels (5-7). Pathological changes that occur in patients with severe SARS-CoV-2 infection, such as increased vascular permeability, local tissue injury and fibrosis, have been attributed to a reduction in mACE2 activity due to binding of SARS-CoV-2 (8, 9).

Angiotensin II stimulates the shedding of soluble ACE2 (sACE2) from mACE2 through a disintegrin and metalloproteinase-17 (ADAM-17) (10). The risk-benefit profile of sACE2 in the context of SARS-CoV-2 infection is disputed. Swärd et al. (2020) proposed that elevated sACE2 levels reflect high mACE2 and/or increased shedding of mACE2 via ADAM-17 and could therefore have value as an indicator of risk for severe SARS-CoV-2 infection (11). Meanwhile, Leow (2020) describes the potential benefits of elevated sACE2 levels, whereby sACE2 could function as a decoy ligand and maintain its affinity for SARS-CoV-2 through the spike protein whilst being unable to mediate cellular entry of the virus (12).

Markers of immune system activation, such as pro-inflammatory cytokine mediators (including interleukin-6 [IL-6]) and proteins of heterogeneous functions that can be activated in pro-inflammatory states (including C-reactive protein [CRP], growth/differentiation factor-15 [GDF-15], and soluble fms-like tyrosine kinase-1 [sFlt-1]), may play a role in SARS-CoV-2 pathology. The cytokine storm in severe COVID-19 is known to be mediated by IL-6 receptors (13-15). Additionally, the acute-phase protein CRP, which is produced by the liver as part of the inflammatory response, has been shown to be a predictive marker of disease progression in patients with severe SARS-CoV-2 infection (16-18) and as an early biomarker for the development of sepsis (19). GDF-15 is part of the transforming growth factor β superfamily of cytokines and elevated levels have been reported in patients with severe SARS-CoV-2 infection (20); GDF-15 has been linked to tissue hypoxia and inflammation (20, 21). In addition, excess levels of the anti-angiogenic protein sFlt-1 have been shown to correlate with endothelial damage and organ failure in patients with SARS-CoV-2 (22).

There is an unmet clinical need for better prediction of disease severity in patients with SARS-CoV-2 to inform patient management and ensure timely treatment. While sACE2 is known to be involved in the pathophysiology of SARS-CoV-2, it is yet to be thoroughly investigated as a biomarker of disease severity. We evaluated the value of sACE2 as a biomarker in the context of other known markers of inflammation and tissue damage (CRP, GDF-15, IL-6, and sFlt-1) in patients with and without SARS-CoV-2 with different clinical outcomes.

## METHODS

### Study design and ethical statement

This was a prospective, observational, single-centre study conducted at Vall d’Hebron University Hospital (Barcelona, Spain) between March and October 2020. The study complied with the principles of the Declaration of Helsinki and received approval (reference: PR [AG] 577/2020) from the Research Ethics Committee for Drug Research of the Vall d’Hebron University Hospital (Barcelona, Spain). Patient-level data were processed in accordance with Regulation (EU) 2016/679 of the European Parliament on Data Protection. An exemption from obtaining patient informed consent was granted by the Research Ethics Committee due to the health emergency presented by the COVID-19 pandemic.

### Samples

Adults aged 25–90 years who were sampled by nasopharyngeal swab for detection of SARS-CoV-2 infection by reverse transcription polymerase chain reaction (RT-PCR) testing (cobas® SARS-CoV-2 real-time RT-PCR test, Roche Diagnostics International Ltd, Rotkreuz, Switzerland) were included in the study. Patients with pathologies related to autoimmune or cardiovascular diseases were excluded; the following comorbidities were not excluded: hypertension, type 2 diabetes mellitus, dyslipidaemia, obesity, and chronic kidney disease.

Serum samples were collected from participants in a random series from March to October 2020 and frozen until analysis. Recruited patients with RT-PCR-confirmed positive SARS-CoV-2 infection at the time of blood draw were grouped according to clinical outcome (**Table 1**): emergency care and home discharge (Group 1); ward admission for moderate illness (Group 2); admission to the intensive care unit (ICU; Group 3); and death associated with SARS-CoV-2 infection (Group 4). Patients who tested negative for SARS-CoV-2 following a previous infection were divided into two groups: an independent cohort of patients who tested negative for SARS-CoV-2 infection one month after finishing a 15-day quarantine following previous RT-PCR-confirmed SARS-CoV-2 infection that required emergency care and home discharge (Group 5) and repeat samples taken from patients in Group 3 previously admitted to the ICU, when they tested negative for SARS-CoV-2 infection immediately before discharge from the hospital (Group 6). Control samples were from adults aged 25–90 years who tested negative for SARS-CoV-2 infection following RT-PCR of a nasopharyngeal swab or had no medical history of SARS-CoV-2 infection (Group 7).

**Table 1.**
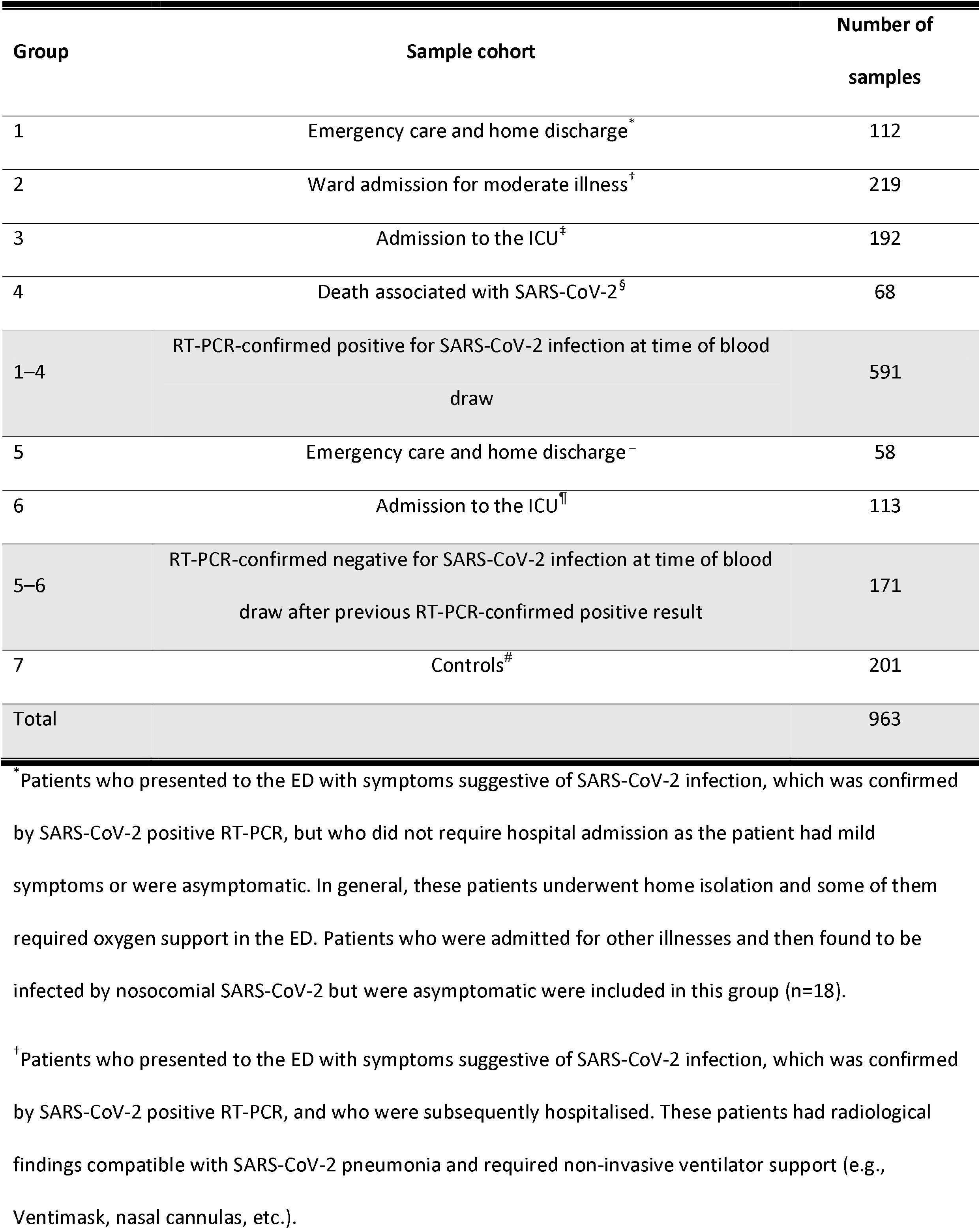
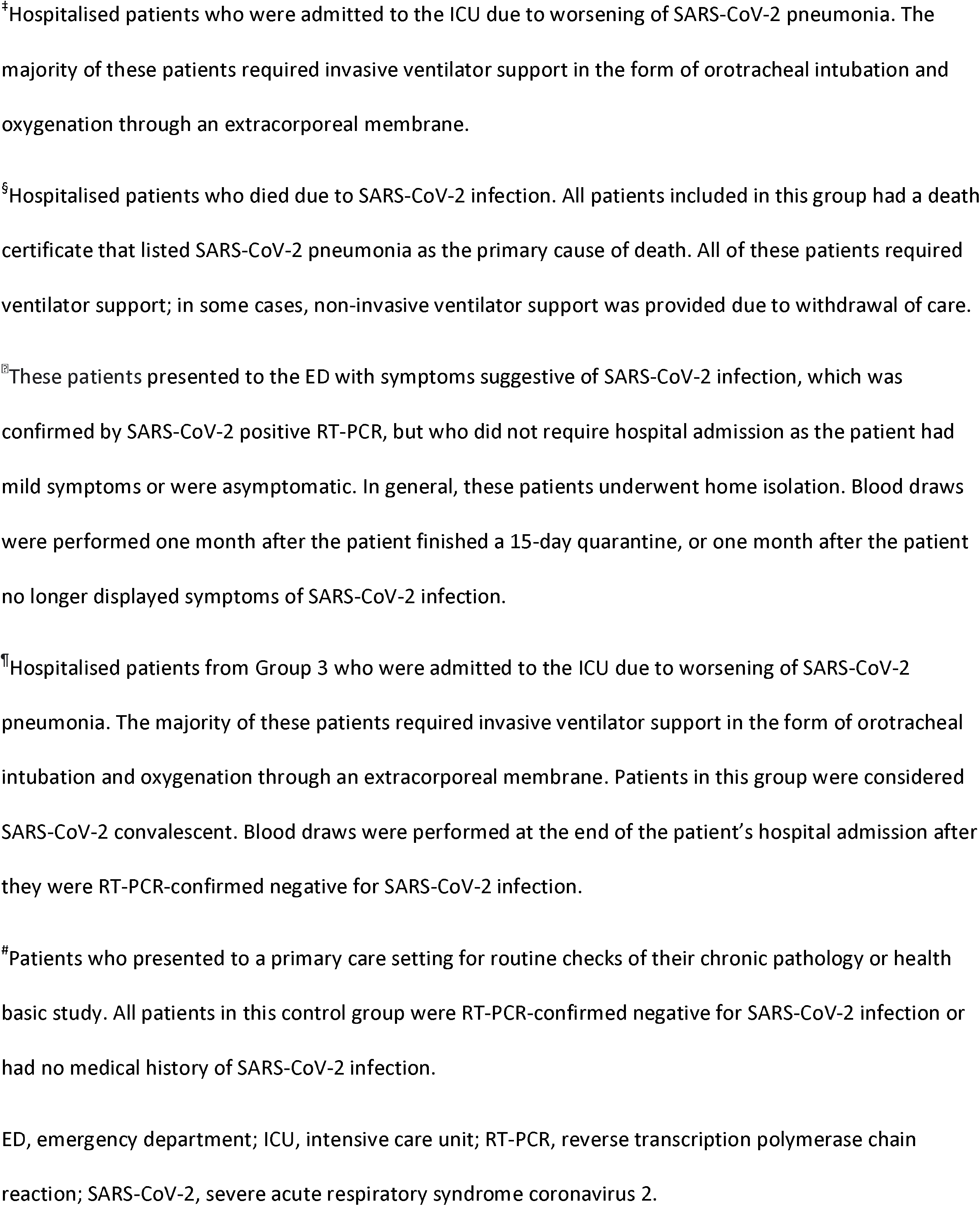
Summary of sample cohorts.

### Sample handling

Anonymised serum samples were thawed and the aliquots stored at 2–8 °C during the week of testing. Following testing, all samples were stored in a serum bank at -80 °C until measurements for all biomarkers were completed. For the control group, serum samples were stored in the serum bank at -80 °C for consistency with the RT-PCR-confirmed SARS-CoV-2 positive serum samples.

### Assays

Serum GDF-15, IL-6, and sFlt-1 levels were measured using the Elecsys® GDF-15, Elecsys IL-6, and Elecsys sFlt-1 electrochemiluminescence immunoassays, respectively, according to the manufacturer’s instructions on the cobas 8000 analyser (all Roche Diagnostics International Ltd). IL-6 was only measured in samples where it had not been previously assessed during routine analysis. Serum CRP levels were measured using an immunoturbidimetry assay on the AU5800 analyser (Beckman Coulter, Brea, CA, US). Serum sACE2 levels were measured using a human ACE2 sandwich enzyme-linked immunosorbent assay (ELISA; Raybiotech, Atlanta, GA, USA) on the Grifols Triturus analyser (Grifols Diagnostic Solutions Inc., Emeryville, CA, USA). Reagents, calibrators, and control materials from the same manufacturer were used for the purpose of this study.

### Data analysis

Sample size was calculated informally according to the estimated number of patients attending the hospital, the service level of the laboratory, and reagent and test availability. Clinical variables for all patients, including demographic data, medical history (including history of liver disease), metabolic profile, and medications were assessed via a review of medical records. Data on the following biochemical variables were also collected: full blood count (including platelets), coagulation (prothrombin time and D-dimer tests), liver function tests (aspartate aminotransferase, alanine aminotransferase, alkaline phosphatase, gamma-glutamyl transferase and bilirubin), lipids (cholesterol and triglycerides), substrates (glucose), renal function (creatinine and urea), and inflammatory markers (IL-6 and CRP). Any missing data was not considered further for statistical analysis.

All values for the statistical analysis were collected, processed, and analysed by researchers at the Vall d’Hebron University Hospital. The data were stored in a pre-specified, anonymised, protected database in electronic format (Excel) and kept at the hospital, where researchers performed the data analysis using R statistical software (version 3.6.2).

The statistical significance of the differences in demographic and biochemical variables was assessed using: Chi-squared test (male sex, chronic kidney disease, blood pressure, type 2 diabetes mellitus, dyslipidaemia, and body mass index), one way ANOVA (age and mean arterial pressure), and Kruskal-Wallis comparison (aspartate aminotransferase, alanine aminotransferase, prothrombin time, and D-dimer). For univariate biomarker analyses, median differences between biomarker levels in patients with SARS-CoV-2 and controls were calculated, and significance determined using the Mann-Whitney U test. Statistical significance was assigned where the p-value was <0.05. Sensitivity and specificity with 95% confidence intervals (CIs) were calculated, and receiver operating characteristic (ROC) analysis used to evaluate area under the curve (AUC). For bivariate biomarker analysis, logistic regression in combination with the mlr R-package (23) was performed. All possible two-biomarker combinations were assessed by means of 10-fold cross-validation; bivariate combinations showing an AUC improvement of at least one percentage point over the best univariate biomarker were reported.

## RESULTS

### Patients

A total of 963 samples from 850 patients were included in the present analyses. A subset of patients had two samples taken: an initial sample that was RT-PCR-confirmed positive for SARS-CoV-2 (Group 3) and a second sample that was negative for SARS-CoV-2 (Group 6). Selected demographic, clinical, and biochemical characteristics of each group are shown in **Table 2**.

**Table 2.** Selected demographic, clinical, and biochemical characteristics of the patient groups.

| Variable | Group |  |  |  |  |  |  | p-value* |
| --- | --- | --- | --- | --- | --- | --- | --- | --- |
|  | RT-PCR-confirmed positive for SARS-CoV-2 |  |  |  | RT-PCR-confirmed negative for SARS-CoV-2 |  |  |  |
|  | 1 | 2 | 3 | 4 | 5 | 6 | 7 |  |
|  | Emergency<br>care &<br>discharge | Ward<br>admission | ICU<br>admission | Death | Emergency<br>care &<br>discharge | ICU admission | Control<br>group |  |
| Age (years), mean (SD) | 59.1 (20.9) | 60.7 (15.4) | 55.4 (11.7) | 71.6 (11.1) | 48.2 (17.3) | 55.2 (11.8) | 61.4 (15.8) | <0.001 |
| Male sex, n (%) | 50 (44.6) | 114 (52.1) | 116 (60.4) | 35 (51.5) | 28 (48.3) | 69 (61.1) | 87 (43.3) | 0.006 |
| Chronic kidney disease<sup>†</sup>, n (%) | 23 (20.5) | 34 (15.6) | 25 (13.1) | 22 (32.4) | 2 (3.6) | 12 (10.6) | 10 (5.0) | <0.001 |
| Arterial pressure (mmHg), mean (SD) | 96.4 (14.2) | 93.7 (12.9) | 91.7 (13.4) | 90.3 (14.9) | 84.6 (11.6) | 92.7 (12.0) | ND | 0.426 |
| Blood pressure >140/90 mmHg, n (%) | 53 (47.3) | 115 (52.5) | 73 (38.0) | 52 (76.5) | 14 (24.1) | 44 (38.9) | 81 (40.3) | <0.001 |
| Type 2 diabetes mellitus, n (%) | 22 (19.6) | 55 (25.1) | 42 (21.9) | 23 (33.8) | 9 (15.5) | 22 (19.5) | 73 (36.3) | 0.001 |
| Dyslipidaemia, n (%)<sup>‡</sup> | 48 (42.9) | 108 (49.3) | 64 (33.3) | 33 (48.5) | 16 (27.6) | 37 (32.7) | 79 (39.3) | 0.003 |
| Body mass index >30 kg/m<sup>2</sup>, n (%) | 28 (25.0) | 79 (36.1) | 64 (33.3) | 18 (26.5) | 5 (8.6) | 38 (33.6) | 37 (18.4) | <0.001 |
| Aspartate aminotransferase (IU/L),<br>median (IQR) | 26.0<br>(21.0, 36.0) | 38.0<br>(29.0, 52.0) | 49.5<br>(31.0, 70.5) | 40.0<br>(28.0, 63.5) | 22.0<br>(19.0, 27.0) | 28.0<br>(22.0, 41.0) | 21.0<br>(18.0, 24.0) | <0.001 |
| Alanine aminotransferase (IU/L),<br>median (IQR) | 20.0<br>(13.0, 30.0) | 27.0<br>(19.0, 50.0) | 40.5<br>(23.0, 65.5) | 24.0<br>(16.0, 36.0) | 19.0<br>(13.0, 28.0) | 45.0<br>(25.0, 60.0) | 18.0<br>(13.0, 24.0) | <0.001 |
| Prothrombin time (INR), median (IQR) | 1.0<br>(1.0, 1.1) | 1.1<br>(1.0, 1.1) | 1.1<br>(1.0, 1.2) | 1.1<br>(1.0, 1.2) | 1.0<br>(0.9, 1.1) | 1.1<br>(1.0, 1.2) | 0.9<br>(0.9, 1.0) | <0.001 |
| D-dimer (ng/mL), median (IQR) | 297.0<br>(155.0,<br>536.0) | 265.0<br>(171.0,<br>432.0) | 391.0<br>(225.0,<br>760.0) | 543.0<br>(216.0,<br>1653.0) | 109.0<br>(50.0, 151.0) | 719.0<br>(307.0, 1567.0) | ND<sup>§</sup> | <0.001 |

\*p-values were calculated using: Chi-squared test (male sex, chronic kidney disease, blood pressure, type 2 diabetes mellitus, dyslipidaemia, and body mass index), one way ANOVA (age and mean arterial pressure), and Kruskal-Wallis comparison (aspartate aminotransferase, alanine aminotransferase, prothrombin time, and D-dimer).
<sup>†</sup>Defined as glomerular filtration rate <60 mL/min/m<sup>2</sup>.
<sup>‡</sup>Defined during routine clinical care, supported by lipid biochemical measurement (cholesterol and triglycerides).
<sup>§</sup>Only one result was available, therefore median and interquartile range could not be calculated.
ANOVA, analysis of variance; ICU, intensive care unit; INR, international normalised ratio; IQR, interquartile range; IU, international units; ND, no data; RT-PCR, reverse transcription polymerase chain reaction; SARS-CoV-2, severe acute respiratory syndrome coronavirus 2; SD, standard deviation.

### Univariate analysis of patients infected with SARS-CoV-2 versus controls

Levels of CRP, GDF-15, IL-6, and sFlt-1 were significantly higher (all p<0.001) in patients infected with SARS-CoV-2 (Groups 1–4) compared with controls (Group 7), whereas levels of sACE2 were significantly lower (p<0.001) in patients infected with SARS-CoV-2 compared with controls (**Table 3**). Based on the analysis of ROC curves, the AUC value (**Table 3**) was highest for CRP (0.964 [95% CI: 0.948, 0.980]), followed by IL-6 (0.949 [95% CI: 0.933, 0.964]), GDF-15 (0.830 [95% CI: 0.797, 0.863]), sFlt-1 (0.797 [95% CI: 0.764, 0.829]), and sACE2 (0.585 [95% CI: 0.539, 0.632]).

**Table 3.** Overview of differences in biomarker levels between patients with SARS-CoV-2 (Groups 1–4) and controls (Group 7).

| Biomarker | Number of samples | | | Biomarker level ($\log_2$),<br>median (IQR) | | | Median<br>difference | p-value | AUC<br>(95% CI) |
| --- | --- | --- | --- | --- | --- | --- | --- | --- | --- |
|  | SARS-CoV-2 | Control | Total | SARS-CoV-2 | Control | Total |  |  |  |
| CRP | 517 | 20 | 537 | 3.26<br>(1.85, 4.14) | -2.08<br>(-3.84, -1.51) | 3.19<br>(1.55, 4.13) | 5.341 | <0.001 | 0.964<br>(0.948, 0.980) |
| GDF-15 | 500 | 201 | 701 | 11.39<br>(10.78, 12.25) | 9.90<br>(9.24, 10.70) | 11.07<br>(10.17, 11.94) | 1.492 | <0.001 | 0.830<br>(0.797, 0.863) |
| IL-6 | 567 | 201 | 768 | 5.56<br>(4.51, 6.64) | 1.21<br>(0.58, 2.06) | 4.90<br>(2.10, 6.22) | 4.355 | <0.001 | 0.949<br>(0.933, 0.964) |
| sACE2 | 591 | 201 | 792 | -4.06<br>(-5.06, -2.00) | -2.84<br>(-5.06, -0.40) | -3.64<br>(-5.06, -1.69) | -1.222 | <0.001 | 0.585<br>(0.539, 0.632) |
| sFlt-1 | 500 | 201 | 701 | 6.88<br>(6.56, 7.18) | 6.45<br>(6.31, 6.58) | 6.69<br>(6.44, 7.05) | 0.431 | <0.001 | 0.797<br>(0.764, 0.829) |

AUC, area under the curve; CI, confidence interval; CRP, C-reactive protein; GDF-15, growth/differentiation factor-15; IL-6, interleukin-6; IQR, interquartile range; sACE2, soluble angiotensin-converting enzyme 2; SARS-CoV-2, severe acute respiratory syndrome coronavirus 2; sFlt-1, soluble fms-like tyrosine kinase-1.

### Univariate and bivariate analysis of patients infected with SARS-CoV-2 who were admitted to hospital versus patients infected with SARS-CoV-2 who were discharged

Levels of CRP, GDF-15, IL-6, and sFlt-1 were significantly higher (all p<0.001) in patients infected with SARS-CoV-2 who were admitted to hospital (Groups 2–4) compared with patients infected with SARS-CoV-2 who were discharged (Group 1), whereas levels of sACE2 were significantly lower (p<0.001) in patients infected with SARS-CoV-2 who were admitted to hospital compared with those who were discharged (**Table 4**). Based on the analysis of ROC curves, the AUC value was highest for IL-6 (0.800 [95% CI: 0.750, 0.851]), followed by CRP (0.775 [95% CI: 0.718, 0.832]), sFlt-1 (0.751 [95% CI: 0.689, 0.813]), sACE2 (0.648 [95% CI: 0.592, 0.704]), and GDF-15 (0.625 [95% CI: 0.551, 0.699]) (**Table 4**). Furthermore, median levels of CRP, GDF-15, IL-6, and sFlt-1 increased with SARS-CoV-2 disease severity, whereas the median level of sACE2 decreased with SARS-CoV-2 disease severity (**Figure 1**).

**Table 4.**
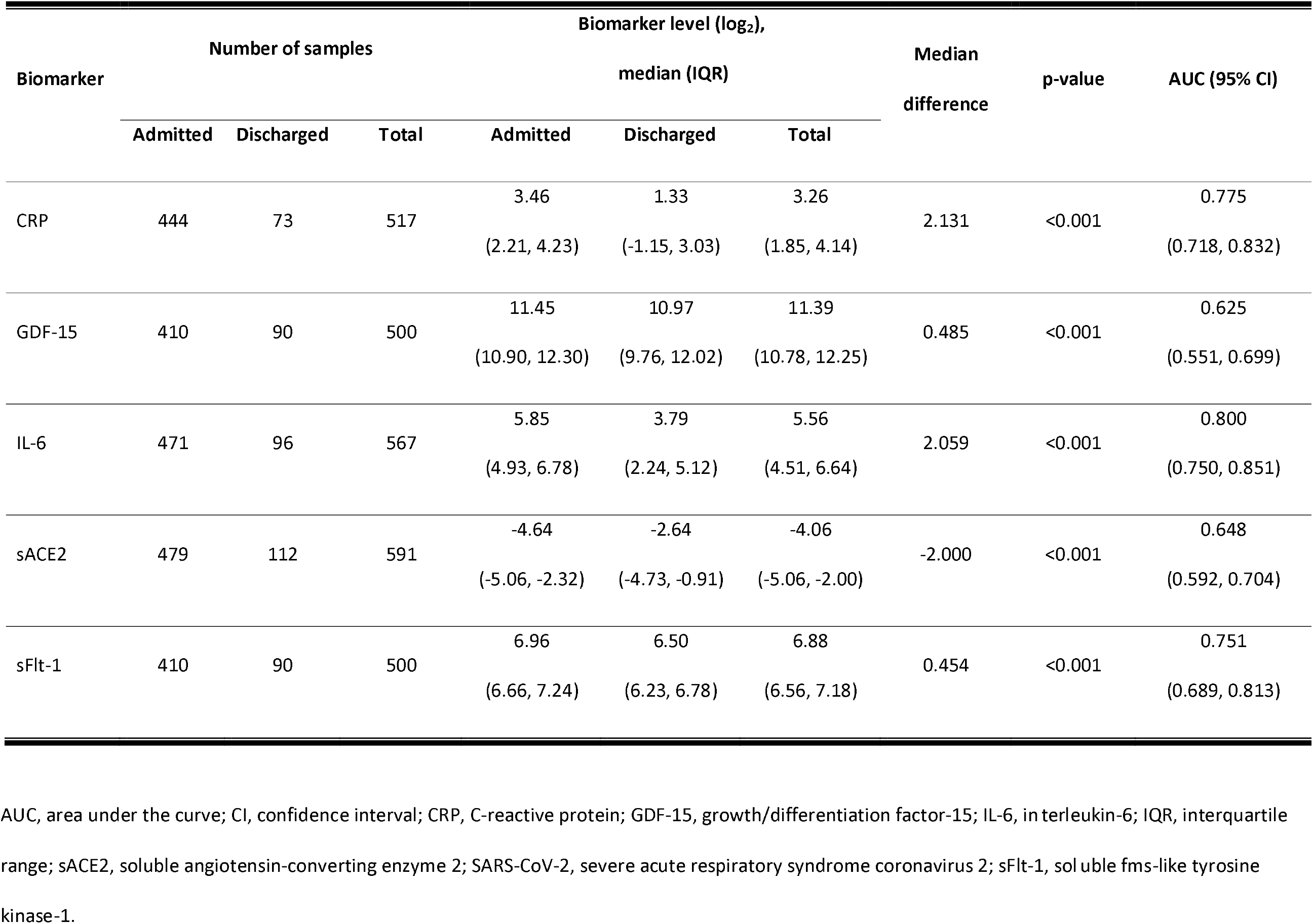
Overview of differences in biomarker levels between patients with SARS-CoV-2 who were admitted to hospital (Groups 2–4) and patients with SARS-CoV-2 who were discharged (Group 1).

| Biomarker | Number of samples | | | Biomarker level ($\log_2$),<br>median (IQR) | | | Median<br>difference | p-value | AUC (95% CI) |
| --- | --- | --- | --- | --- | --- | --- | --- | --- | --- |
|  | Admitted | Discharged | Total | Admitted | Discharged | Total |  |  |  |
| CRP | 444 | 73 | 517 | 3.46<br>(2.21, 4.23) | 1.33<br>(-1.15, 3.03) | 3.26<br>(1.85, 4.14) | 2.131 | <0.001 | 0.775<br>(0.718, 0.832) |
| GDF-15 | 410 | 90 | 500 | 11.45<br>(10.90, 12.30) | 10.97<br>(9.76, 12.02) | 11.39<br>(10.78, 12.25) | 0.485 | <0.001 | 0.625<br>(0.551, 0.699) |
| IL-6 | 471 | 96 | 567 | 5.85<br>(4.93, 6.78) | 3.79<br>(2.24, 5.12) | 5.56<br>(4.51, 6.64) | 2.059 | <0.001 | 0.800<br>(0.750, 0.851) |
| sACE2 | 479 | 112 | 591 | -4.64<br>(-5.06, -2.32) | -2.64<br>(-4.73, -0.91) | -4.06<br>(-5.06, -2.00) | -2.000 | <0.001 | 0.648<br>(0.592, 0.704) |
| sFlt-1 | 410 | 90 | 500 | 6.96<br>(6.66, 7.24) | 6.50<br>(6.23, 6.78) | 6.88<br>(6.56, 7.18) | 0.454 | <0.001 | 0.751<br>(0.689, 0.813) |

AUC, area under the curve; CI, confidence interval; CRP, C-reactive protein; GDF-15, growth/differentiation factor-15; IL-6, interleukin-6; IQR, interquartile range; sACE2, soluble angiotensin-converting enzyme 2; SARS-CoV-2, severe acute respiratory syndrome coronavirus 2; sFlt-1, soluble fms-like tyrosine kinase-1.

**Figure 1.**
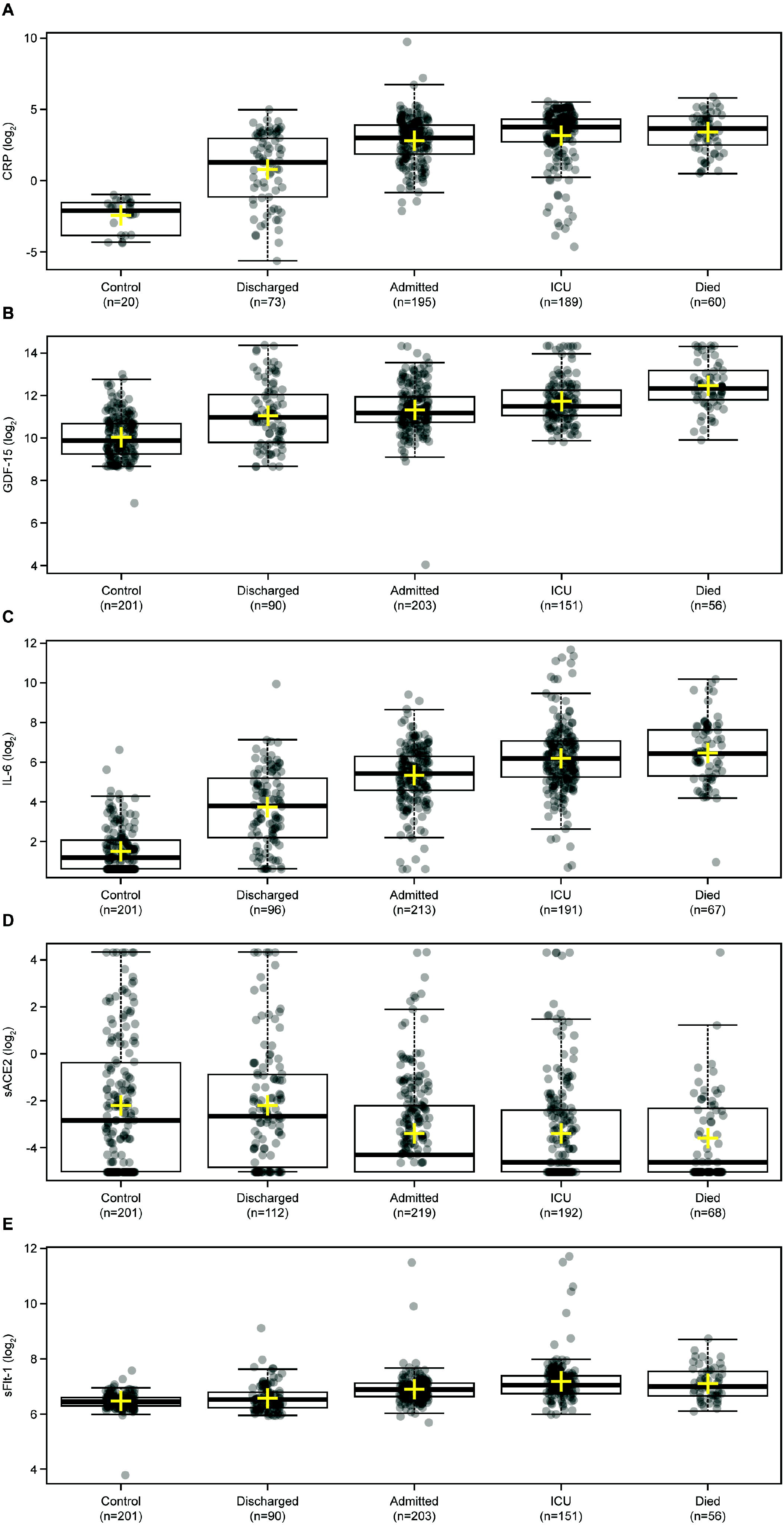
Boxplots displaying the levels of (A) CRP, (B) GDF-15, (C) IL-6, (D) sACE2, and (E) sFlt-1 according to disease severity in patients with SARS-CoV-2 who were admitted to hospital (Groups 2–4) and patients with SARS-CoV-2 who were discharged (Group 1). The thick black line indicates the median and the yellow cross indicates the mean. The boxplot upper/lower limits and whiskers denote the interquartile range and maximum/minimum values excluding outliers (i.e., values beyond 1.5 times the interquartile range from the box), respectively. CRP, C-reactive protein; GDF-15, growth/differentiation factor-15; ICU, intensive care unit; IL-6, interleukin-6; sACE2, soluble angiotensin-converting enzyme 2; SARS-CoV-2, severe acute respiratory syndrome coronavirus 2; sFlt-1, soluble fms-like tyrosine kinase-1.

Bivariate analysis based on ROC curves (**Figure 2**) showed that the addition of sFlt-1 (cross-validated [cv] AUC=0.832), GDF-15 (cvAUC=0.832) or sACE2 (cvAUC=0.812) to IL-6 provided an improvement in AUC value compared with IL-6 alone (AUC: 0.800) in patients with SARS-CoV-2 who were admitted to hospital (Groups 2–4) versus patients with SARS-CoV-2 who were discharged (Group 1).

**Figure 2.**
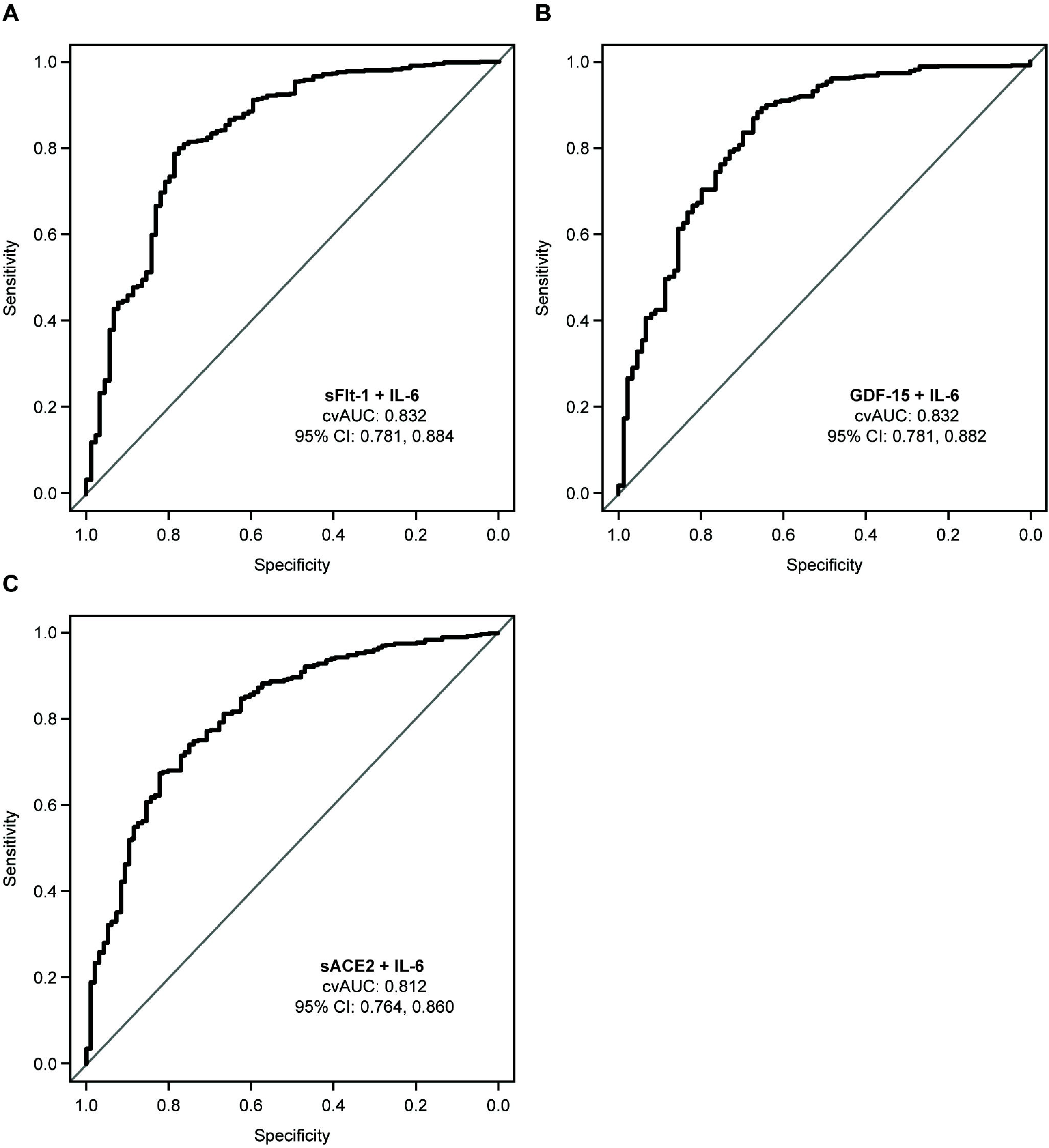
Bivariate analysis performed in patients with SARS-CoV-2 who were admitted to hospital (Groups 2–4) versus patients with SARS-CoV-2 who were discharged (Group 1). Data are shown as ROC curves for (A) sFlt-1 + IL-6 (cvAUC: 0.832), (B) GDF-15 + IL-6 (cvAUC: 0.832), and (C) sACE2 + IL-6 (cvAUC: 0.812). cvAUC, cross-validated area under the curve; CI, confidence interval; GDF-15, growth/differentiation factor-15; IL-6, interleukin-6; ROC, receiver operating characteristic; sACE2, soluble angiotensin-converting enzyme 2; SARS-CoV-2, severe acute respiratory syndrome coronavirus 2.

As sACE2 has not previously been explored as a biomarker for SARS-CoV-2 disease severity, two cut-offs were proposed for sACE2 based on ROC curve analysis for the prediction of hospitalisation versus discharge in patients infected with SARS-CoV-2 (**Figure 3**). A cut-off of ≤0.05 ng/mL produced a positive predictive value (PPV) of 89.1% (95% CI: 84.9, 92.5) based on a disease prevalence of 81% and a cut-off of ≥0.42 ng/mL provided a negative predictive value (NPV) of 84.0% (95% CI: 80.4, 87.1) based on a disease prevalence of 19%.

**Figure 3.**
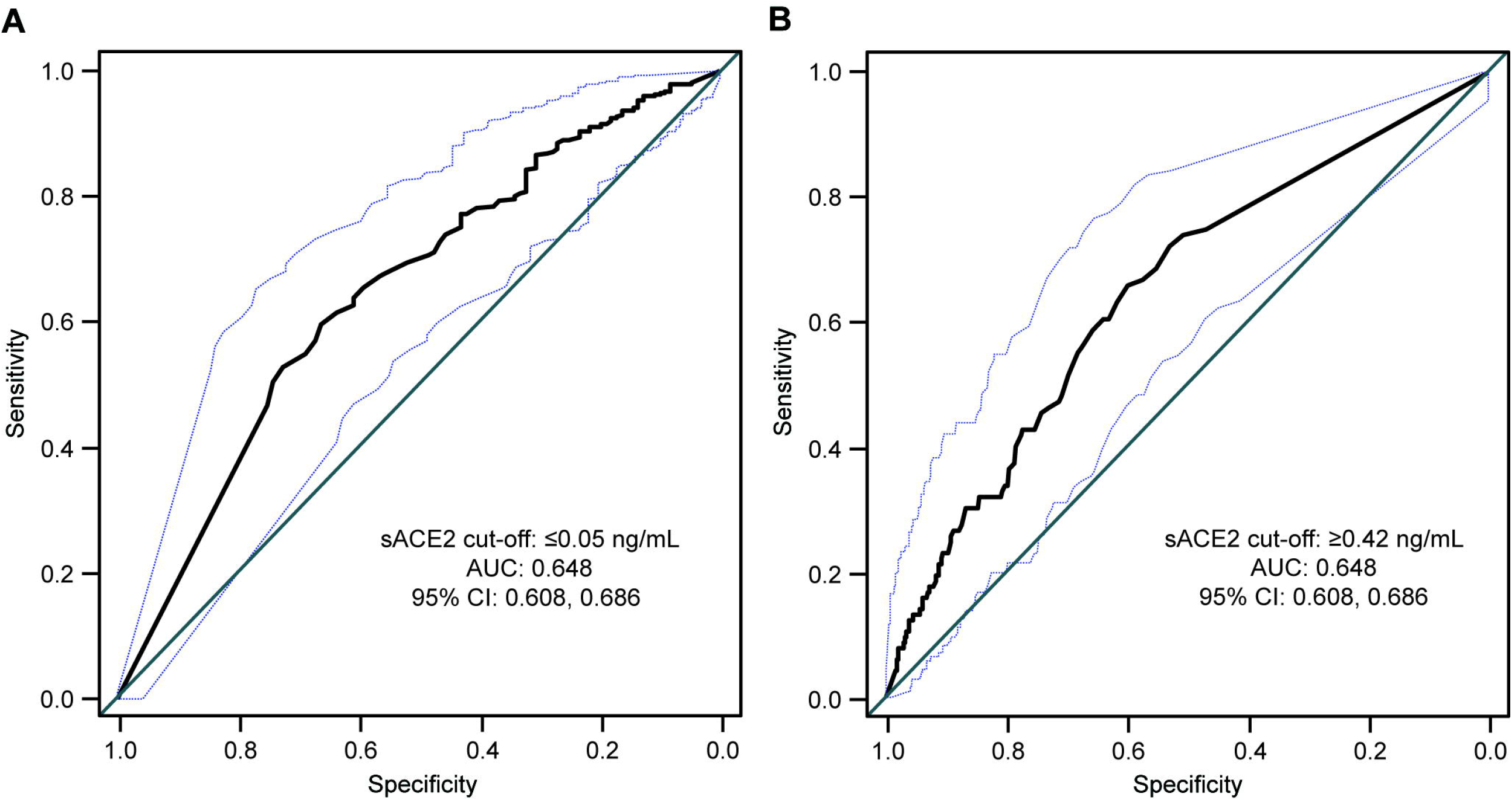
ROC curve analysis used to generate cut-off values of sACE2 for the prediction of hospitalisation in patients with SARS-CoV-2. A cut-off of ≤0.05 ng/mL (A) produced a positive predictive value of 89.1% (95% CI: 84.9, 92.5) where prevalence of disease was 81% and a cut-off of ≥0.42 ng/mL (B) provided a negative predictive value of 84.0% (95% CI: 80.4, 87.1) where prevalence of disease was 19%, for the prediction of hospitalisation. The AUC for both panels A and B was 0.648 (95% CI: 0.608, 0.686). The blue dashed lines indicate the upper and lower limits of the 95% CI for AUC. AUC, area under the curve; CI, confidence interval; ROC, receiver operating characteristic; sACE2, soluble angiotensin-converting enzyme 2; SARS-CoV-2, severe acute respiratory syndrome coronavirus 2.

### Univariate analysis of patients infected with SARS-CoV-2 who were admitted to the ICU or died versus patients infected with SARS-CoV-2 who were admitted to the ward or discharged

Levels of CRP, GDF-15, IL-6, and sFlt-1 were significantly higher (all p<0.001) in patients infected with SARS-CoV-2 who were admitted to the ICU or died (Groups 3 and 4) compared with patients infected with SARS-CoV-2 who were admitted to the ward or discharged (Groups 1 and 2; **Table 5**). sACE2 levels were significantly lower (p=0.015) in patients who were admitted to the ICU or died, compared with patients who were admitted to the ward or discharged. Based on the analysis of ROC curves, the AUC value was highest for IL-6 (0.715 [95% CI: 0.673, 0.757]), followed by sFlt-1 (0.672 [95% CI: 0.624, 0.720]), CRP (0.670 [95% CI: 0.623, 0.716]), GDF-15 (0.650 [95% CI: 0.602, 0.698]), and sACE2 (0.556 [95% CI: 0.511, 0.600]).

**Table 5.** Overview of differences in biomarker levels between patients with SARS-CoV-2 who were admitted to the ICU or died (Groups 3 and 4) and patients with SARS-CoV-2 who were admitted to the ward or discharged (Groups 1 and 2).

| Biomarker | Number of samples | | | Biomarker level ($\log_2$),<br>median (IQR) | | | Median<br>difference | p-value | AUC (95% CI) |
| --- | --- | --- | --- | --- | --- | --- | --- | --- | --- |
|  | ICU or<br>died | Ward or<br>discharged | Total | ICU or died | Ward or<br>discharged | Total |  |  |  |
| CRP | 249 | 268 | 517 | 3.74<br>(2.70, 4.42) | 2.77<br>(1.32, 3.71) | 3.26<br>(1.85, 4.14) | 0.969 | <0.001 | 0.670<br>(0.623, 0.716) |
| GDF-15 | 207 | 293 | 500 | 11.74<br>(11.09, 12.46) | 11.15<br>(10.54, 11.94) | 11.39<br>(10.78, 12.25) | 0.593 | <0.001 | 0.650<br>(0.602, 0.698) |
| IL-6 | 258 | 309 | 567 | 6.19<br>(5.21, 7.10) | 5.12<br>(3.85, 6.05) | 5.56<br>(4.51, 6.64) | 1.073 | <0.001 | 0.715<br>(0.673, 0.757) |
| sACE2 | 260 | 331 | 591 | -4.64<br>(-5.06, -2.40) | -3.64<br>(-5.06, -1.71) | -4.06<br>(-5.06, -2.00) | -1.000 | 0.015 | 0.556<br>(0.511, 0.600) |
| sFlt-1 | 207 | 293 | 500 | 7.02<br>(6.71, 7.38) | 6.77<br>(6.49, 7.05) | 6.88<br>(6.56, 7.18) | 0.252 | <0.001 | 0.672<br>(0.624, 0.720) |

AUC, area under the curve; CI, confidence interval; CRP, C-reactive protein; GDF-15, growth/differentiation factor-15; ICU, intensive care unit; IL-6, interleukin-6; IQR, interquartile range; sACE2, soluble angiotensin-converting enzyme 2; SARS-CoV-2, severe acute respiratory syndrome coronavirus 2; sFlt-1, soluble fms-like tyrosine kinase-1.

### Patients who were RT-PCR-confirmed negative for SARS-CoV-2 infection following a previous positive RT-PCR result

Median sACE2 levels were similar (p=0.273) between samples from patients who were RT-PCR-confirmed positive for SARS-CoV-2 at the time of blood draw and received emergency care then discharged at the time of infection (Group 1; 0.160 ng/mL) and samples from a different group of patients who received similar care who were RT-PCR-confirmed negative for SARS-CoV-2 infection at the time of blood draw (Group 5; 0.235 ng/mL). In contrast, Median sACE2 levels were significantly higher (p=0.003) in samples from patients who were RT-PCR-confirmed negative for SARS-CoV-2 infection following a previous positive RT-PCR result and admitted to the ICU (Group 6; 0.130 ng/mL), relative to samples taken from the same patients when they were RT-PCR-confirmed positive for SARS-CoV-2 (Group 3; 0.040 ng/mL).

In all patients who were RT-PCR-confirmed negative for SARS-CoV-2 infection following a positive RT-PCR result (Groups 5 and 6), a significant increase (p=0.049) in sACE2 levels was observed in those who were discharged (median sACE2 level: 0.235 ng/mL) compared with those who were admitted to the ICU (median sACE2 level: 0.130 ng/mL).

## DISCUSSION

We evaluated the value of sACE2 as a novel biomarker for disease severity in the context of other markers of inflammation and tissue damage (CRP, GDF-15, IL-6, and sFlt-1) in a large cohort of patients with and without SARS-CoV-2 with different clinical outcomes. Univariate analysis showed that median levels of sACE2 were significantly lower in patients infected with SARS-CoV-2 compared with controls, in patients infected with SARS-CoV-2 who were admitted to hospital compared with patients infected with SARS-CoV-2 who were discharged, and in patients infected with SARS-CoV-2 who were admitted to the ICU or who died compared with patients who were discharged or admitted to the ward. These findings suggest that sACE2 has value as a biomarker for hospitalisation and disease severity in patients with SARS-CoV-2. Furthermore, two cut-offs for sACE2 for predicting severe SARS-CoV-2 infection were derived: ≤0.05 ng/mL with a PPV of 89.1% and ≥0.42 ng/mL with a NPV of 84.0%. After future validation in larger studies in an acute care setting, the cut-off of ≤0.05 ng/mL could be used to indicate risk of severe disease when IL-6 or CRP values are not elevated to sufficient levels due to very recent SARS-CoV-2 infection. Meanwhile, a cut-off of ≥0.42 ng/mL could help identify the majority of patients who develop mild disease.

CRP had the highest AUC value when comparing patients infected with SARS-CoV-2 versus controls, and IL-6 had the highest AUC when comparing patients infected with SARS-CoV-2 who were admitted to hospital versus those who were discharged and when comparing patients infected with SARS-CoV-2 who were admitted to the ICU or died versus those who were admitted to the ward or discharged. Overall, median levels of CRP, GDF-15, IL-6 and sFlt-1 were significantly higher in patients with SARS-CoV-2 who had the most severe clinical outcome in all comparisons. These findings are consistent with previously published studies that have reported the value of these biomarkers for disease severity in patients infected with SARS-CoV-2 (16, 20, 22, 24).

Across all patients with SARS-CoV-2, sACE2 levels were significantly lower than in controls, which could indicate a reduced amount of mACE2 susceptible to cleavage by ADAM-17. In addition, the median level of sACE2 was significantly lower in patients infected with SARS-CoV-2 with the most severe clinical outcome in all comparisons. These findings support the hypothesis that sACE2 could play a protective role in patients infected with SARS-CoV-2 (12). One mechanism through which sACE2 could have this protective effect is by remaining residually active and performing negative feedback in the activation of RAS. In keeping with this, human recombinant sACE2 has been explored as a treatment option for patients infected with SARS-CoV-2 and was associated with a decrease in concentration of critical cytokines implicated in SARS-CoV-2 pathology, reductions in the viral load, and neutralisation of viral particles; however, speculation remains whether these observations were reflective of the sACE2 treatment or the natural course of the viral infection (25, 26).

In contrast to our findings, a recent study by Kragstrup et al. reported that high plasma ACE2 was associated with increased maximal illness severity within 28 days; however, the patient population in this study was different to the population in the current study in that it did not examine longitudinal samples from the same patient during hospitalisation. In addition, plasma ACE2 was measured using a different methodology (protein extension assay), which does not allow the determination of a cut-off value for prediction of severe disease and limits the comparability of findings with other studies (27). Another study in a small number of patients reported increased sACE2 in patients with severe SARS-CoV-2 infection (28); however, this increase has been hypothesised to be a transient response due to increased shedding from infected cells (29). Thus, the timing of sampling from a positive SARS-CoV-2 RT-PCR test may be an important factor in interpreting sACE2 levels. It is also important to note that sACE2 is expressed in other organs in addition to the lungs (30), and a blood draw is indicative of universal circulating levels of sACE2. Local sACE2 levels may be a more accurate indicator of SARS-CoV-2 severity and explain the differences in response to infection, degree of severity, and recovery period observed between patients. Circulating levels of sACE2 have also been shown to vary by gender and age (11, 29).

In patients infected with SARS-CoV-2 who were admitted to the ICU, levels of sACE2 were significantly higher in blood samples drawn following the resolution of the infection (0.130 ng/mL) compared with blood samples drawn at the time of RT-PCR-confirmed positivity for SARS-CoV-2 (0.040 ng/mL), which suggests that levels of sACE2 are able to rise following infection. In further support of a protective role of sACE2, patients infected with SARS-CoV-2 who were admitted to the ICU had approximately half of the sACE2 levels compared with patients infected with SARS-CoV-2 who were discharged in samples taken after the viral infection had resolved (0.130 ng/mL versus 0.235 ng/mL).

One strength of this study is the large number of samples from different patient groups that were used to determine median biomarker levels. In addition, the observed age range of all individuals included in this study (48.2–71.6 years) was reflective of the group most likely to be at risk of more severe clinical outcomes when infected with SARS-CoV-2, relative to younger patients (31). A limitation of this study is the single-centre design and under-representation of certain groups (i.e., patients infected with SARS-CoV-2 who were discharged and controls). In addition, the patients recruited with mild symptoms of SARS-CoV-2 infection were relatively complicated in terms of their clinical presentation, as they were recruited from the emergency department. It was not possible to obtain samples from patients with mild symptoms that did not require hospital treatment due to the public health recommendation to stay at home during the period of this study. Additionally, sACE2 measurements in this study were performed using a non-commercial kit for which the quality specifications are not as robust as those of the other markers used in clinical practice.

Future validation of the present biomarker measurements in an independent cohort of patients is warranted. In particular, longitudinal studies of sACE2 levels in patients who have recovered from severe SARS-CoV-2 infection are required to confirm if sACE2 levels increase after a total recovery of symptoms or if they are permanently reduced following severe SARS-CoV-2 infection. Furthermore, characterisation of sACE2 levels is required in a younger population, in individuals with and without hypertension and chronic kidney disease, and in patients receiving treatments that may affect sACE2 activity or concentration (i.e., ACE inhibitors or angiotensin receptor blockers).

In conclusion, the present findings support the further investigation of sACE2 as a novel biomarker for hospitalisation risk and disease severity in patients infected with SARS-CoV-2, which could be used to complement and improve the performance of other biomarkers of inflammation and tissue damage.

## Data Availability

The study was conducted in accordance with applicable regulations. Ethical approval was provided by the Research Ethics Committee for Drug Research of the Vall d'Hebron University Hospital (Barcelona, Spain). An exemption from obtaining patient informed consent was granted by the Research Ethics Committee due to the health emergency presented by the COVID-19 pandemic. Therefore, participants of this study did not give consent for their data to be shared with additional third parties. For more information on the study and data sharing, qualified researchers may contact the corresponding author Francisco Rodriguez-Frias.

## ACKNOWLEDGEMENTS

Third-party medical writing support for the development of this manuscript, under the direction of the authors, was provided by Heather Small, PhD, of Ashfield MedComms (Macclesfield, UK), an Ashfield Health company, and was funded by Roche Diagnostics International Ltd (Rotkreuz, Switzerland). COBAS and ELECSYS are trademarks of Roche. All other product names and trademarks are the property of their respective owners.

